# Multiomic prioritisation of risk genes for anorexia nervosa

**DOI:** 10.1101/2022.06.04.22275898

**Authors:** Danielle M. Adams, William R. Reay, Murray J. Cairns

## Abstract

Anorexia nervosa is the leading cause of mortality among psychiatric disorders worldwide. Currently no medications are approved for anorexia nervosa treatment, and thus, identification of risk factors for this disorder is pivotal to improve patient outcomes. We used models of genetically imputed expression and splicing from 17 tissues (two blood and fifteen brain regions), leveraging mRNA, protein, and mRNA alternative splicing weights to identify genes, proteins, and transcripts, respectively, associated with anorexia nervosa risk. We uncovered 134 genes for which genetically predicted mRNA expression was associated with anorexia nervosa after multiple-testing correction, as well as four proteins and 16 alternatively spliced transcripts. Conditional analysis of these significantly associated genes on other proximal association signals resulted in 97 genes independently associated with anorexia nervosa. Moreover, probabilistic finemapping further refined these associations and prioritised putative causal genes. The gene *WDR6*, for which increased genetically predicted mRNA expression was correlated with anorexia, was strongly supported by both conditional analyses and finemapping. Pathway analysis of genes revealed by finemapping identified regulation of *immune system process* (overlapping genes = *MST1, TREX1, PRKAR2A, PROS1*) as statistically overrepresented. In summary, we leveraged multiomic datasets to genetically prioritise novel risk genes for anorexia nervosa that warrant further exploration.

## INTRODUCTION

Anorexia nervosa is a complex psychiatric disorder associated with alterations to satiety, activity, and self-perception that results in severe mental distress and malnourishment (1). Anorexia nervosa usually begins during adolescence with a long course of illness and one of the highest mortality rates of any psychiatric disorder (2). Cognitive behavioural therapy and weight rehabilitation are the first-line treatments for anorexia nervosa, as there are still no approved pharmacotherapies for the disorder (3). Unravelling the biological complexity of anorexia onset and its clinical course will be key to developing more effective interventions and improving clinical management.

Anorexia is a complex disorder influenced by genetic and environmental factors, with twin studies estimating heritability at 56% (4), and common variants are now shown to account for around 20% of this through genome-wide association studies (GWAS) (5). The most recent anorexia nervosa GWAS uncovered eight independent genome-wide associated loci (6). While this presents an opportunity to identify new drug targets, many of the causal variants in these loci will be confounded by linkage disequilibrium (LD) and other interactions that obscure their effects on specific genes.

Gene-based aggregation methods can increase power to detect associations beyond genome-wide significant loci and yield more biologically relevant information. For example, transcriptome wide association studies (TWAS) achieve this by integrating mRNA expression data with GWAS association data to detect genes for which genetically predicted expression is associated with the trait (7). This approach can be extended to other quantitative functional data such as protein and alternatively spliced mRNA isoform abundance. TWAS is capable of prioritising genes that may be involved in the disorder and also assign them a direction of expression that is odds increasing. (7). Despite these useful features TWAS alone is not a test of causality as the signal does not exclude the possibility of co-regulation between genes or LD between variants used in expression models (8). Interrogating TWAS results through approaches such as conditional analysis and probabilistic finemapping can be utilised to overcome some of these limitations and identify genes from association signals that could constitute a causal relationship (9, 10).

Anorexia TWAS data have previously been leveraged to reveal insights into the pathogenesis of the disorder (11-13), however, these studies did not specifically prioritise genes that could be involved in the disorder. In addition, extending TWAS to the use of protein and mRNA isoform weights has not previously been investigated for anorexia nervosa and could further identify novel risk genes. In this study, we utilise models of genetically predicted mRNA expression, protein expression, and alternative mRNA isoform abundance to prioritise genes involved in anorexia nervosa. These associations signals were further refined through conditional analysis and finemapping to reveal several genes including WDR6 with plausible utility as targets for therapeutic intervention. Pathways analysis also implicated regulation of *immune system process* with gene set functional analysis including signal from *MST1, TREX1, PRKAR2A* and *PROS1*.

## METHODS

### Overview of study

We aimed to prioritise genes associated with anorexia nervosa and implicate potential mechanisms related to expression and/or splicing. Firstly, we conducted a comprehensive brain and blood-based transcriptome-wide association study (TWAS) (8, 14, 15). TWAS requires expression data which can be imputed from independent SNP-mRNA expression weights from multivariate models of *cis*-acting genetically regulated expression (GReX). TWAS then compares imputed expression with SNP-anorexia effect sizes to test the association between predicted expression and the odds of anorexia. Genes uncovered from this approach that survived multiple-testing correction were then further probed to refine candidate causal genes through conditional analysis and probabilistic fine-mapping (10, 14). Whilst mRNA expression is arguably the most well studied cellular readout, genes may operate more specifically in the pathogenesis of anorexia nervosa through dysregulation of other factors like protein expression and alternative splicing. As a result, we leveraged SNP weights, where available, for protein expression and splicing to also perform a proteome-wide association study (PWAS) and an alternative splicing based test (spliceWAS). Genes were prioritised based on evidence from mRNA, protein, and alternative isoform expression and then subjected to further *in silico* analyses related to overrepresentation in biological pathways, spatiotemporal expression, and *in vivo* phenotypes. The anorexia nervosa GWAS utilised in this study was a meta-analysis European ancestry cohort that totalled 16,992 cases and 55,525 controls – further details related to collection of samples, phenotype acquisition, and GWAS approach are described in the original publication (6).

### Weights for genetically predicted mRNA, protein, and splicing

Brain related SNP weights (multivariate GReX) for TWAS were derived from GTEx v7 and PsychENCODE, whilst whole blood weights were also obtained from GTEx v7 (14, 16). The GTEx v7 SNP weights comprise data from twelve different brain regions – with the sample sizes of the cohorts utilised for GReX estimation as follows: amygdala (N=88), anterior cingulate cortex (N=109), caudate (N=144), cerebellar hemisphere (N=125), cerebellum (N=154), cortex (N=136), frontal cortex (N=136), hippocampus (N=111), hypothalamus (N=108), nucleus accumbens (N=130), putamen (N=80) and substantia nigra (N=80). HapMap3 SNPs from the 1000 genomes phase 3 European reference panel were used as a linkage disequilibrium (LD) estimate, to correspond with how the weights were calculated with those same HapMap3 SNPs. The TWAS using whole blood weights utilised the same LD reference strategy, with 369 GTEx v7 participants in these models. Frontal or cerebral cortex tissue from the larger PsychENCODE cohort was also utilised in terms of SNP weights, with a sample size of 1695. As described in the original PsychENCODE publication, gene-wise GReX were estimated for all imputed SNPs, not just the HapMap3 panel, and thus, we utilised the full suite of the phase 3 1000 genomes European subset as the LD reference. There were two tissues for which SNP weights related to protein expression were available – the dorsolateral prefrontal cortex (DLPFC, N = 376) and plasma (N = 7213) (17, 18). It should be noted that the plasma weights were derived from the European subset of the study cohort and the authors only used the elastic net method to derive GReX. Analogous to the difference between the GTEx v7 and PsychENCODE studies above, the DLPFC SNP weights were estimated using the HapMap3 panel, and thus, we only used those SNPs as an LD reference. The plasma PWAS utilised the full reference panel. Finally, the splicing related weights were derived from the DLPFC samples from the Common Mind Consortium (N=452) which utilised the HapMap3 restricted 1000 genomes panel (19).

### Implementation of the FUSION pipeline

We conducted TWAS/PWAS/spliceWAS using the FUSION package (14). Specifically, the *FUSION*.*assoc_test*.*R* script was utilised, with the best performing GReX model selected from five-fold cross-validation (*R*^2^) and the SNP weight set selected by FUSION for calculating the TWAS *Z* score. In accordance with usual practice for the FUSION approach, only genes/transcripts with significantly non-zero *cis*-acting heritability (*cis*-*h*^2^) are included. Prior to analysis, summary statistics were also munged whereby SNPs were retained with an imputation INFO > 0.9, as well as removing indels, strand ambiguous SNPs and SNPs with MAF < 0.01. We corrected for the number of non-missing SNP weight sets using the Bonferroni [family-wise error rate (FWER) based] and Benjamini-Hochberg [false discovery rate (FDR) based] approaches for the TWAS, PWAS, and spliceWAS independently.

### Conditional analyses and probabilistic fine-mapping

We applied conditional analysis via the FUSION framework (*FUSION*.*post_process*.*R*) to the regions of significant genes after correction to investigate the proportion of implicated genes in any given locus that are independently associated. Specifically, jointly significant genes retain their significance after jointly estimating association for all models within a 500,000 base pair region of a significant gene. Marginally associated genes, which are not jointly significant, likely arise due to factors such as genes for which predicted expression was correlated.

Moreover, we applied probabilistic finemapping to prioritise candidate causal genes from any locus in the anorexia GWAS summary statistics with at least a suggestively significant SNP (*P* < 1 × 10^−5^) using the FOCUS method (10). The default prior (*p* = 1 × 10^−3^) and prior variance (*nσ*^2^ = 40) were utilised to approximate Bayes’ factors such that the posterior inclusion probability (*PIP*) of each gene being a member of a credible set with 90% probability of containing the causal gene could be derived. Finemapping was performed with default tissue prioritisation in addition to the prioritisation of brain tissue. In the TWAS, there were two reference panels utilised for fine-mapping – for genes uncovered from a GTEx v7 tissue, we utilised the default combined FOCUS SNP weight set which collated GTEx v7 tissues, DLPFC (CommonMind), blood (YFS, NTR), and adipose (METSIM) SNP weight sets (https://www.dropbox.com/s/ep3dzlqnp7p8e5j/focus.db?dl=0), with genes discovered using the PsychENCODE weights fine-mapped specifically using that panel given the different LD parameters and its more complete set of genes with *cis-*heritable models in that one tissue. The multi-tissue fine-mapping panel contains several other non-brain tissues, and thus, some GReX models that would not have been available in brain and blood. We sought to balance maximising the number of models available for finemapping, whilst acknowledging that some of the tissues in this panel are less likely to be disease relevant. As FOCUS allows the null mode that the causal feature is not typed to be predicted as a possible member of the credible set, we excluded any genes for which that occurred. The credible set was defined by summing normalised *PIP* such that *ρ* was exceeded, sorting the genes, and then including those genes until at least *ρ* of the normalised-posterior mass is explained, as described in more detail elsewhere (10, 20). As an exploratory analysis, we also applied finemapping to the PWAS and spliceWAS results, although the limited number of SNP weights available for protein and alternative splicing means that the probability of a causal gene for any region not being present is higher.

### Investigation of prioritised anorexia nervosa associated genes

We further investigated two sets of prioritised genes *in silico*: 1) conditionally independent associations (TWAS/PWAS/spliceWAS) from marginally significant signals (FDR < 0.05) that are at least nominally jointly significant (*P* < 0.05), and 2) genes in the fine-mapped 90% credible set with *PIP* > 0.4 and the absence of the null model in the credible set. Firstly, we considered biological pathways and other ontological sets for which these two sets of genes could separately be overrepresented via the g:Profiler framework and the Benjamini-Hochberg method for multiple-testing correction (21). Moreover, the GENE2FUNC module of FUMA (functional mapping and annotation of genetic associations) was utilised to test for tissue specific expression amongst different tissues (GTEx v8: 54 tissue types) and temporal stages of brain development (BrainSpan: 29 different ages of brain samples) (22). Interactions between proteins may indicate shared mechanisms of action, and thus, protein-protein interactions for these gene-sets were estimated using STRING (23). Protein-protein interactions were obtained from experiment and database sources (interaction score > 0.7) using a 5% false discovery rate. STRING estimates the interactions between proteins within a set, in addition to quantifying if the observed number of interactions is greater than expected given the size of the gene-set.

## RESULTS

### Novel anorexia risk genes uncovered through genetically predicted expression or splicing

Firstly, a transcriptome-wide association study was performed using models of genetically predicted mRNA expression across twelve brain regions from GTEx, cortical samples from the PsychENCODE consortium, and whole blood GTEx samples. This was the most well-powered approach as there were 57,596 mRNA models available to test, with the number of unique genes totalling 11,242, 12,183, and 5,915, for GTEx brain, PsychENCODE brain, and GTEx whole blood samples, respectively (*Supplementary Table 1*). The most significant association signals were found in a gene dense region on chromosome 3 (*Figure 1)*, in line with expectation given the significant GWAS signal for anorexia nervosa (chromosome 3: 47,588,253-51,368,253) overlaps this cluster of significant genes. Upregulation of the gene encoding WD Repeat Domain 6 (*WDR6*) was the top hit in this region associated with increased odds of anorexia nervosa – *Z*_TWAS_ = 7.44, *P* = 9.96 × 10^−14^ (GTEx cortex). This gene also surpassed Bonferroni correction in several other tissues, including the larger sample size PsychENCODE cortical samples, whole blood, nucleus accumbens, and caudate basal ganglia. The WD repeat protein family effects signal transduction (24), whilst *WDR6* is thought to influence cell cycle arrest (25). Several other proximal genes to *WDR6* also survived Bonferroni correction in multiple tissues, such as, *CCDC71, NCKIPSD*, and *MST1R. O*-6-Methylguanine-DNA Methyltransferase (*MGMT*) was the most significant gene outside of chromosome 3 (*Z*_TWAS_ = -5.33, *P* = 9.88 × 10^−8^ – caudate basal ganglia), followed by *CDK11B* on chromosome 1 and the long non-coding RNA (lncRNA) *LINC00324* on chromosome 17. Previous analysis, differing in methodology and samples found an association between *SUOX* expression and anorexia which was not replicated in the present study (12, 26).

**Figure 1:**
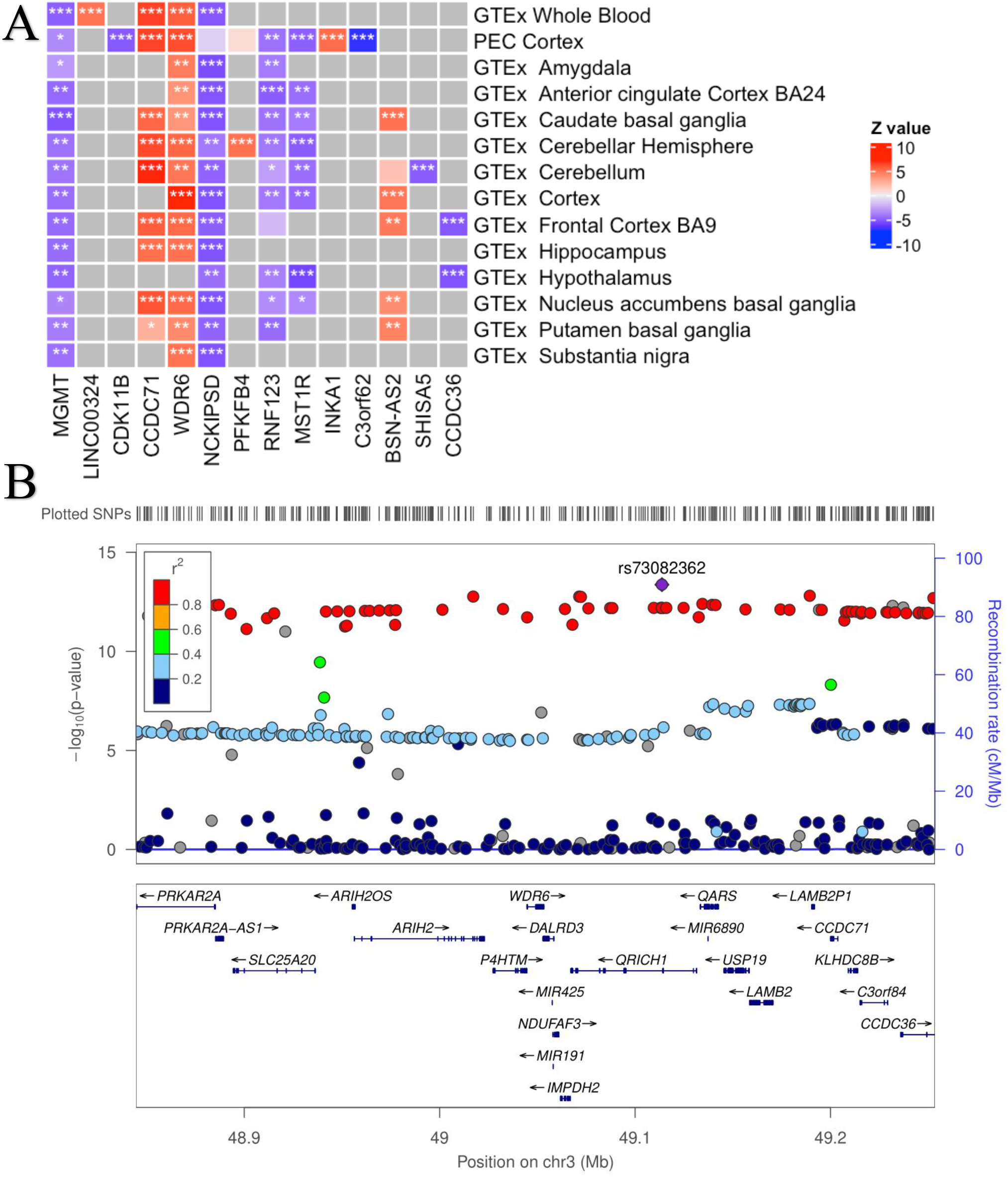
TWAS associations and region plot of the densely associated anorexia signal on chromosome 3. A: Heatmap of genes with at least one Bonferroni significant eQTL tissue associated with anorexia nervosa. Red indicates positive z scores, blue indicates negative z scores (legend). Columns indicates genes, rows indicate tissue models. * Indicates nominally significant genes, ** indicates Benjamini-Hochberg significant association, *** indicates Bonferroni significant association. Grey squares indicate that a significantly cis-heritable model of imputed expression data was unavailable in that tissue. B: Relative anorexia nervosa gene and SNP locations and significance. Points in the top panel indicate SNPs, legend indicates r^2^, left side y-axis indicates the negative log transformed p-value of SNPs, right side y-axis indicates the recombination rate (cM/Mb). The bottom panel indicates the location of genes relative to the top panel SNPs. Plot generated using ZoomLocus (31) with 200kb flanking size. The SNP with the most significant p-value in this region: rs73082362 is highlighted.

We then sought to extend the power for gene discovery, as well as supporting signals uncovered by the TWAS, through integration of models of genetically regulated protein expression (PWAS) and alternative splicing (spliceWAS, *Supplementary Tables 2, 3*). There were 2,300 protein expression SNP weight sets in total available to test across the DLPFC and blood cohorts in which the imputed expression models were trained. Two proteins survived Bonferroni correction in the PWAS, and we uncovered four signals that were significant after applying a less stringent FDR based cut-off (FDR < 0.05). The strongest protein association, MHC class I chain-related gene B (MICB) (*Z*_PWAS_ = 4.53, P= 5.75×10^−6^, plasma) was correlated with increased odds risk of anorexia, however, due to the extensive LD of the MHC region it is difficult to apply approaches such as PWAS, and thus, this signal should be treated cautiously. The densely associated region on chromosome 3 harboured the next most significant protein expression signal, with decreased predicted expression of Glutathione peroxidase 1 (GPX1) associated with anorexia in the DLPFC (*Z*_PWAS_ = -4.41), mirroring its negative TWAS test statistic in the hypothalamus for mRNA expression (*Z*_TWAS_ = -4.51). Thereafter, the remaining two proteins surviving correction were each on different chromosomes (*FGF23*, chr 12 and *CTNND1*, chr 11). The spliceWAS yielded fourteen transcripts that survived Bonferroni correction from the 7,708 transcripts tested relating to 3,291 total genes. The mean number of alternatively spliced transcripts per gene that were significantly *cis*-heritable and available for analyses was 2.3. Once more the anorexia nervosa association dense region on chromosome 3 implicated already by GWAS, TWAS, and PWAS, yielded the most significant signal, with seven transcripts of *ARIH2* significantly associated. Interestingly, there were mixed directions of effect amongst the different isoforms as increased predicted abundance of three splice variants were associated with anorexia, whilst the converse was true for the remaining four. None of these isoforms correspond to the canonical transcript; chr3:48918821:48967151 (27). In general, the *ARIH2* gene is postulated to regulate ubiquitination (28, 29) and post-transcriptionally modifies *NLRP3* to reduce inflammatory activity (30), however, the functional specificity of particular splicing isoforms is less well characterised. This gene was also not significant in the TWAS, despite having a well-powered model of imputed expression in the PsychENCODE cortical samples. The only remaining transcripts surviving Bonferroni correction not proximally located to the chromosome 3 region were two isoforms of GPR75-ASB3 Readthrough (*GPR75-ASB3*), which are isoforms from which a naturally occurring readthrough occurs between two genes (*GPR75* and *ASB3*), and thus, features exons from both genes. Decreased predicted abundance of these two isoforms was associated with anorexia.

We then intersected the TWAS, PWAS, and spliceWAS results, finding that of the genes modelled, or a transcript thereof, 631 were available with a significantly *cis*-heritable model for all three modalities (mRNA, protein, alternative splicing), no genes significantly (Benjamini-Hochberg) overlapped all three methods. From the 134 Benjamini-Hochberg significant TWAS genes, four of these were nominally significant (*P* < 0.05) for protein expression, while 13 were nominally significant for isoform expression. Two of the Four Benjamini-Hochberg significant PWAS genes were nominally significant for TWAS without any isoform overlap. Of the 16 Benjamini-Hochberg significantly associated gene isoforms from the spliceWAS, 12 had nominally significant mRNA expression models, while only one had a nominally significant protein expression model. The *Z* scores from models with available data across all three modalities exhibited low correlation (mRNA-protein: *r* = 0.07 [*P*=1.3×10^−11^], protein-isoform: *r* = -0.008 [*P*=0.49], mRNA-isoform: *r* = 0.02 [*P*=0.04]), however, the correlation between mRNA and protein expression was statistically significant.

### Conditional analyses and probabilistic fine-mapping further refine anorexia association signals

Co-regulation and LD between genes can confound TWAS signals and lead to spurious associations for non-causal genes. Conditional analysis and finemapping was performed to distinguish genes with increased evidence of exerting an independent causal effect on anorexia nervosa. Conditional analysis (14) estimates the residual independent association of TWAS signals after controlling for the predicted expression of nearby significant genes. Benjamini-Hochberg significant TWAS genes were subjected to conditional analysis to predict which genes accounted for the localised signal. From 313 significant TWAS signals, 97 genes had a conditionally independent association (*P*_Joint_ < 0.05) with anorexia nervosa as indicated by their nominally significant joint *P* value (*Supplementary table 4*). The gene most significantly associated with anorexia, *WDR6* (*Z*_conditional-TWAS_=7.4, *P*=1×10^−13^, Cortex) maintains an independent association after conditioning on the 77 other TWAS significant gene models (21 genes) from chromosome 3, three of which are also conditionally independent (*CTNNB1, GOLIM4, STX19*).

Finemapping (10) is a Bayesian statistical method designed to isolate subsets of genes more likely to contain causal genes. We applied this approach to all suggestively significant regions in the anorexia GWAS (*P*_GWAS_ < 1 × 10^−5^) to derive 90% credible sets. Credible sets were removed if they contained the null model that is included by FOCUS to account for missing causal mechanisms like genes without a suitable GReX model. Across either mRNA, protein or mRNA alternative splicing weights there were 116 genes that were prioritised in a credible set (*Supplementary table 5*). Of these eight genes (*Table* 1) had moderate (*PIP* > 0.4) evidence of a causal effect on anorexia, while five genes demonstrated strong evidence (*PIP* > 0.8). The strongest evidence of a causal relationship was observed for Neurexophilin And PC-Esterase Domain Family Member 1 (*NXPE1*) (*PIP*=1, testis). Surprisingly, *NXPE1* was indicated for the testis (GTEx), whilst it was also highly expressed in the colon (32). The other four genes with strong evidence of a potential causal effect on anorexia were as follows: *WDR6* (*PIP*= 0.997, DLPFC), *PRKAR2A* (*PIP*= 0.814, GTEx artery tibial), *PROS1 (PIP*= 0.895, GTEx cortex) and the non-coding RNA *RP13-238F13*.*5 (PIP*= 0.971, GTEx spinal cord cervical c-1). *WDR6* and *MST1* were the only genes found to be both conditionally independent with at least moderate finemap evidence of a causal relationship (*Table 1*). Macrophage Stimulating 1 (*MST1*) mediates cell division and apoptosis (33, 34) and is predicted to increase risk of anorexia (*Z*_TWAS_ = 4.85, P= 1.2×10^−6^, GTEx Hypothalamus). However, finemapping of the MST1 PWAS indicates evidence of a protective relationship with anorexia (*Z*_finemap_ = -4.63, PIP=0.457, blood plasma protein). This discordant direction between mRNA and protein requires further investigation to refine its biological salience. The power of finemapping to identify causal genes increases when more GReX expression models are included, and thus, tissues were used for finemapping that were not subjected to the marginal TWAS, since removing these would decrease finemapping power. There are three genes (*RP13-28F13*.*5, NXPE1* and *PRKAR2A*) without TWAS blood and brain associations. One of these; *PRKAR2A* exists in a credible set with four other genes (the same set as the fine-mapped *TREX1*), three of which are associated within brain regions. The DGIdb drug bank was queried to determine if any FDA approved pharmaceutical interventions exist which target these anorexia nervosa associated genes. The DGIdb drug bank indicated six drugs which target *PROS1* (35) a fine-mapped gene for which increased genetically predicted expression was associated with increased anorexia nervosa risk. One of these drugs, menadione is indicated to activate *PROS1* (36, 37), however, as a risk increasing gene it would need to be inhibited to potentially treat anorexia nervosa. There are currently no approved drugs for the other fine-mapped genes (*MST1, WDR6, RP13-238F13*.*5, NXPE1, C3orf62, TREX1* and *PRKAR2A*) which could be an avenue for future research.

**Table 1:** Finemap associations. Finemapped genes with moderate or strong evidence of a causal effect on anorexia (PIP>0.4). Tissue indicates which tissue the expression weights were derived from, Ref name indicates the group/consortium responsible for eQTL generation, Z_finemap_ indicates the estimated Z score association, PIP indicates the posterior inclusion probability and region indicates the GRCh37 genomic region the credible set was derived from. Credible set size indicates the number of genes in the credible set when prioritising brain or any tissue. All genes are in the credible set. Z_TWAS_ (P) -tissue and Z_PWAS_ (P) -tissue indicates the TWAS and PWAS Z score and P value associated with the most significant tissue for each association. No spliceWAS associations are available for these eight genes.

| Molecular name | tissue | Ref name | $Z_{finemap}$ | PIP | $Z_{TWAS}(P)$ -tissue | $Z_{PWAS}(P)$ -tissue | region | Credible set size (brain/default) |
| --- | --- | --- | --- | --- | --- | --- | --- | --- |
| *MST1* | Blood plasma protein | gtex | -4.63 | 0.457 | 4.85 ($1.2 \times 10^{-6}$) - hypothalamus - Conditionally independent | -3.4787 ($5.04 \times 10^{-4}$) - plasma | 3:49317941-3:51830833 | NA/1 |
| *WDR6* | brain dorsolateral prefrontal cortex | pec | 6.46 | 0.997 | 7.44 ($9.96 \times 10^{-14}$) - cortex - Conditionally independent | NA | 3:49317338-3:51828635 | 1/1 |
| *RP13-238F13.5* | brain spinal cord cervical c-1 | gtex | 5.01 | 0.97 | NA | NA | 10:125919795-10:127999424 | 1/1 |
| *NXPE1* | testis | gtex | 6.02 | 1 | NA | NA | 11:114833710-11:116383064 | 1/1 |
| *C3orf62* | brain dorsolateral prefrontal cortex | cmc | -7.19 | 0.423 | -7.13 ($9.71 \times 10^{-13}$) - cortex | NA | 3:47777774-3:49313978 | 4/384 |
| *TREX1* | brain cortex | gtex | -6.95 | 0.449 | -3.2575 ($1.1 \times 10^{-3}$) - Nucleus accumbens basal ganglia | -2.1349 (0.032) - DLPFC | 3:49317338-3:51815687 | 5/476 |
| *PROS1* | brain cortex | gtex | 5.21 | 0.895 | 0.07 (0.94) - whole blood | NA | 3:94256654-3:95306175 | 2/3 |
| *PRKAR2A* | artery tibial | gtex | -6.95 | 0.814 | NA | NA | 3:49317338-3:51815687 | 5/476 |

### Functional interrogation of prioritised anorexia nervosa risk genes suggests a role for immune function

Pathway analysis was performed on the conditionally independent (N = 97) and fine-mapped credible set (N = 8) genes using g:Profiler (Ensembl 103, Ensembl Genomes 50). We focused on pathways that were enriched for either of these input sets of genes that survived FDR correction (FDR < 0.05) and had at least three intersecting genes with the input list. Firstly, the conditionally independent genes were overrepresented amongst pathways related to the *presynaptic active zone* (overlapping genes = *STX19, CTNND1, CTBP2, CTNNB1*) (*Supplementary Table 6*). While the genes prioritised by fine-mapping were overrepresented in the *regulation of immune system process* (overlapping genes = *MST1, TREX1, PRKAR2A, PROS1*) (*Supplementary Table 7*).

To determine the potential functional mechanisms through which finemapped and conditionally independent genes may affect anorexia nervosa, we investigated tissue specificity, temporal expression and protein interactions. Both the predicted conditionally independent genes and the finemapped credible genes with moderate evidence of a causal effect on anorexia were investigated with STRING and FUMA (*Supplementary figure 1-4*). STRING (*Supplementary figure 5, 6*) indicated that fewer protein-protein interactions occur within the set of finemapped genes (STRING-*P*=1) and conditionally independent genes (STRING-*P*=0.4) than expected. This may indicate that there are no biological mechanism linking the finemapped genes, however these results may also be explained by incomplete availability of data. Considering anorexia nervosa onset predominately occurs during adolescence it would be expected that genes that impact anorexia aetiology may be differentially expressed during this period, however FUMA GENE2FUNC did not indicate any consistent increased or decreased differential expression during adolescence. Similarly, there was no systemic patterns of tissue expression across these sets of genes, however it is not a prerequisite for genes to exhibit differential expression in the regions with which they exert an effect. These methods are limited by the accuracy and depth of gene functional data which may prevent the analysis of pathways or genes that are poorly characterised.

## DISCUSSION

Anorexia nervosa is a complex disorder characterised by weight loss, altered self-perception and other comorbidities such as anxiety. To advance new treatments we need to increase our understanding of the pathogenic mechanisms that underlie the onset and progression of the disorder, as cognitive behavioural therapy is the only approved treatment (3). In the current study we integrated genome wide association signal for anorexia nervosa with genetically regulated gene expression to identify potential drugs targets for the disorder.

Triangulation of evidence across statistical association methods enhances the confidence in the disorder relevance of a gene. Upregulation of WD Repeat Domain 6 (*WDR6*) was supported in our study as exhibiting a plausible causal relationship with anorexia nervosa across all methods with available data (TWAS, conditional analysis, fine-mapping). The region surrounding *WDR6* is dense with genes also predicted to have an association with anorexia nervosa but due to linkage disequilibrium, refining causal genes is challenging. A conditionally significant TWAS signal along with the finemapping evidence suggests that *WDR6* is a causal gene at this locus, however, further analyses such as SNP based finemapping and *in silico* prediction of variants in this locus are warranted in future to confirm this finding given genetic influence on disease may not directly be mediated by *cis*-acting expression. *WDR6* is a conserved repeat region expression ubiquitously during human development that may function as a restriction factor to inhibit virus replication and a protein complex assembly platform (38-40). In rats *WDR6* physically interacts with insulin receptor substrate 4 to modulate insulin signalling in the hypothalamic arcuate nucleus (41). Both WDR6 (42) and circulating insulin (43) inhibit autophagy. While incidence of apoptosis and autophagy particularly in the liver are increased in anorexia patients (44, 45). Additionally, fasting insulin level has evidence to suggest it exerts a protective effect against anorexia nervosa risk (46) which may relate to the predicted effect of *WDR6* on anorexia nervosa.

Accumulating observational evidence suggests a relationship may exist between immune function and anorexia nervosa (47, 48). Starvation can lead to inflammation; however, there remains differences between the presentation of immune dysregulation during malnutrition and anorexia nervosa, suggesting an underlying relationship may exist beyond the pathology associated with malnutrition (49). Interestingly, four of the eight fine-mapped genes including *MST1, TREX1, PRKAR2A*, and *PROS1* were a member in the *immune system processes* gene-set, which was a statistically significant overrepresentation. This is supportive of previous observations suggesting that immune system dysregulation may contribute to anorexia nervosa onset and maintenance (49). Furthermore, the strongest protein PWAS signal MICB, which implicates the MHC region, and the lead mRNA isoform gene *ARIH2* were also present within the immune system processes pathway. However, the overrepresentation in this instance was not significant like the finemapped genes. The identification of an MHC protein (MICB) in the PWAS necessitates further analysis of the MHC region in anorexia nervosa, such as the role of specific human leukocyte antigen (HLA) types. Conditionally independent or finemapped genes from this pathway; *MST1* (50, 51), *PRKAR2A* (52) and *ARIH2* (30) have been linked to inflammatory processes in previous studies. Anorexia nervosa is observationally associated with a pro-inflammatory state which includes increased levels of cytokines such as tumour necrosis factor and interleukins 1 and 6, in addition to T cell proliferation (49, 53, 54). Although these studies are likely confounded by factors like reverse causality that complicates their interpretation. Recent genetic evidence from our group demonstrates evidence of a protective effect of C-reactive protein (CRP) on anorexia nervosa, which may relate to infection susceptibility given that CRP is not simply a marker of inflammation, as is often characterised, and directly participates in processes like phagocytosis (48, 55). Our data suggests that genes with immune related functions are involved in the pathogenesis of anorexia nervosa. Further work is now needed to refine whether the immune system is a plausible target for anorexia nervosa that could lead to new treatments.

Whilst TWAS, SpliceWAS and PWAS provide a mechanistic framework for the associative evidence between genes and disease, there is also significant confounding by co-regulation derived correlation. TWAS associations can also be confounded by linkage disequilibrium which can bias SNP effect estimates for both expression weights and disease associations (7). The performance of TWAS is also limited to some extent by the sample size of GReX data from different tissues and that some genes are not expressed and are therefore “missing” from a relevant gene set. Incompleteness of gene expression data diminishes the effectiveness of finemapping, null models within the credible set could be better linked to potentially causal genes which would allow for the identification of other causal genes. Analyses performed in exclusively European ancestries will not have the benefit of variant diversity than can support the finemapping and when larger and more diverse non-European samples and trans-ancestry GWAS become available, this is likely to improve TWAS and fine mapping studies. Another limitation in the existing anorexia nervosa GWAS has been the predominately female composition which restricts the identification of sex specific causal risk/protective factors. Increased male case acquisition and diagnosis for future GWAS is therefore likely to improve risk characterisation across demographics.

In conclusion, our study highlighted novel genes, proteins and mRNA isoforms predicted to effect anorexia risk. In particular, we provide strong evidence suggesting that *WDR6* and several genes related to immune system function contribute to the pathogenesis of the disorder. Further research is warranted to establish these mechanisms and determine their potential as targets for treatment.

## Data Availability

All data in this study are publicly available. GWAS summary statistics for anorexia nervosa are available to download from the Psychiatric Genomics Consortium (https://www.med.unc.edu/pgc/download-results/). The TWAS and spliceWAS SNP weights are available from (http://gusevlab.org/projects/fusion/), whilst the PWAS SNP weights can be obtained from (http://nilanjanchatterjeelab.org/pwas/) and (https://www.synapse.org/#!Synapse:syn23627957) for plasma and brain, respectively.

http://gusevlab.org/projects/fusion/

https://www.synapse.org/#!Synapse:syn23627957

## ACKNOWLEDGEMENTS

This work was supported by a National Health and Medical Research Council (NHMRC) grant (1188493). M.J.C. is supported by an NHMRC Senior Research Fellowship (1121474).The funders had no role in study design, data collection and analysis, decision to publish, or preparation of the paper. The authors wish to acknowledge the Eating Disorders working group of the Psychiatric Genomics Consortium for making their summary statistics publicly available.

## AUTHOR CONTRIBUTIONS

W.R.R. conceived and designed the study with input from D.M.A and M.J.C. D.M.A performed the analyses. D.M.A, W.R.R. and M.J.C wrote the manuscript. W.R.R. and M.J.C contributed to the interpretation and visualisation of results.

## REFERENCES

1. Sibeoni J, Orri M, Valentin M, Podlipski M-A, Colin S, Pradere J, et al. Metasynthesis of the Views about Treatment of Anorexia Nervosa in Adolescents: Perspectives of Adolescents, Parents, and Professionals. PLOS ONE. 2017;12(1):e0169493.

2. Edakubo S, Fushimi K. Mortality and risk assessment for anorexia nervosa in acute-care hospitals: a nationwide administrative database analysis. BMC Psychiatry. 2020;20(1):19.

3. Wonderlich SA, Bulik CM, Schmidt U, Steiger H, Hoek HW. Severe and enduring anorexia nervosa: Update and observations about the current clinical reality. International Journal of Eating Disorders. 2020;53(8):1303–12.

4. Bulik CM, Sullivan PF, Tozzi F, Furberg H, Lichtenstein P, Pedersen NL. Prevalence, Heritability, and Prospective Risk Factors for Anorexia Nervosa. Archives of General Psychiatry. 2006;63(3):305–12.

5. Hirtz R, Hinney A. Genetic and epigenetic findings in anorexia nervosa. Medizinische Genetik. 2020;32(1):25–9.

6. Watson HJ, Yilmaz Z, Thornton LM, Hübel C, Coleman JRI, Gaspar HA, et al. Genome-wide association study identifies eight risk loci and implicates metabo-psychiatric origins for anorexia nervosa. Nat Genet. 2019;51(8):1207–14.

7. Wainberg M, Sinnott-Armstrong N, Mancuso N, Barbeira AN, Knowles DA, Golan D, et al. Opportunities and challenges for transcriptome-wide association studies. Nat Genet. 2019;51(4):592–9.

8. Reay WR, Cairns MJ. Advancing the use of genome-wide association studies for drug repurposing. Nature Reviews Genetics. 2021;22(10):658–71.

9. Hall LS, Medway CW, Pain O, Pardiñas AF, Rees EG, Escott-Price V, et al. A transcriptome-wide association study implicates specific pre-and post-synaptic abnormalities in schizophrenia. Hum Mol Genet. 2019;29(1):159–67.

10. Mancuso N, Freund MK, Johnson R, Shi H, Kichaev G, Gusev A, et al. Probabilistic fine-mapping of transcriptome-wide association studies. Nat Genet. 2019;51(4):675–82.

11. Cheng B, Qi X, Liang C, Zhang L, Ma M, Li P, et al. Integrative Genomic Enrichment Analysis Identified the Brain Regions and Development Stages Related to Anorexia Nervosa and Obsessive-Compulsive Disorder. Cereb Cortex. 2020;30(12):6481–9.

12. Chatzinakos C, Georgiadis F, Lee D, Cai N, Vladimirov VI, Docherty A, et al. TWAS pathway method greatly enhances the number of leads for uncovering the molecular underpinnings of psychiatric disorders. Am J Med Genet B Neuropsychiatr Genet. 2020;183(8):454–63.

13. Johnson JS, Cote AC, Dobbyn A, Sloofman LG, Xu J, Cotter L, et al. Mapping anorexia nervosa genes to clinical phenotypes. Psychol Med. 2022:1–15.

14. Gusev A, Ko A, Shi H, Bhatia G, Chung W, Penninx BW, et al. Integrative approaches for large-scale transcriptome-wide association studies. Nat Genet. 2016;48(3):245–52.

15. Gamazon ER, Wheeler HE, Shah KP, Mozaffari SV, Aquino-Michaels K, Carroll RJ, et al. A gene-based association method for mapping traits using reference transcriptome data. Nat Genet. 2015;47(9):1091–8.

16. Gandal MJ, Zhang P, Hadjimichael E, Walker RL, Chen C, Liu S, et al. Transcriptome-wide isoform-level dysregulation in ASD, schizophrenia, and bipolar disorder. Science. 2018;362(6420):eaat8127.

17. Wingo AP, Liu Y, Gerasimov ES, Gockley J, Logsdon BA, Duong DM, et al. Integrating human brain proteomes with genome-wide association data implicates new proteins in Alzheimer’s disease pathogenesis. Nat Genet. 2021;53(2):143–6.

18. Zhang J, Dutta D, Köttgen A, Tin A, Schlosser P, Grams ME, et al. Large Bi-Ethnic Study of Plasma Proteome Leads to Comprehensive Mapping of <em>cis</em>-pQTL and Models for Proteome-wide Association Studies. bioRxiv. 2021:2021.03.15.435533.

19. Gusev A, Mancuso N, Won H, Kousi M, Finucane HK, Reshef Y, et al. Transcriptome-wide association study of schizophrenia and chromatin activity yields mechanistic disease insights. Nat Genet. 2018;50(4):538–48.

20. Reay WR, El Shair SI, Geaghan MP, Riveros C, Holliday EG, McEvoy MA, et al. Genetic association and causal inference converge on hyperglycaemia as a modifiable factor to improve lung function. Elife. 2021;10.

21. Reimand J, Kull M, Peterson H, Hansen J, Vilo J. g:Profiler--a web-based toolset for functional profiling of gene lists from large-scale experiments. Nucleic Acids Res. 2007;35(Web Server issue):W193–200.

22. Watanabe K, Taskesen E, van Bochoven A, Posthuma D. Functional mapping and annotation of genetic associations with FUMA. Nature Communications. 2017;8(1):1826.

23. Szklarczyk D, Gable AL, Lyon D, Junge A, Wyder S, Huerta-Cepas J, et al. STRING v11: protein-protein association networks with increased coverage, supporting functional discovery in genome-wide experimental datasets. Nucleic Acids Res. 2019;47(D1):D607–D13.

24. Li D, Roberts R. Human Genome and Diseases:¶WD-repeat proteins: structure characteristics, biological function, and their involvement in human diseases. Cellular and Molecular Life Sciences CMLS. 2001;58(14):2085–97.

25. Xie X, Wang Z, Chen Y. Association of LKB1 with a WD-repeat protein WDR6 is implicated in cell growth arrest and p27(Kip1) induction. Mol Cell Biochem. 2007;301(1-2):115–22.

26. Baird DA, Liu JZ, Zheng J, Sieberts SK, Perumal T, Elsworth B, et al. Identifying drug targets for neurological and psychiatric disease via genetics and the brain transcriptome. PLoS Genet. 2021;17(1):e1009224.

27. Howe KL, Achuthan P, Allen J, Allen J, Alvarez-Jarreta J, Amode MR, et al. Ensembl 2021. Nucleic Acids Research. 2020;49(D1):D884–D91.

28. Kelsall IR, Duda DM, Olszewski JL, Hofmann K, Knebel A, Langevin F, et al. TRIAD1 and HHARI bind to and are activated by distinct neddylated Cullin-RING ligase complexes. Embo j. 2013;32(21):2848–60.

29. Marteijn JA, van Emst L, Erpelinck-Verschueren CA, Nikoloski G, Menke A, de Witte T, et al. The E3 ubiquitin-protein ligase Triad1 inhibits clonogenic growth of primary myeloid progenitor cells. Blood. 2005;106(13):4114–23.

30. Kawashima A, Karasawa T, Tago K, Kimura H, Kamata R, Usui-Kawanishi F, et al. ARIH2 Ubiquitinates NLRP3 and Negatively Regulates NLRP3 Inflammasome Activation in Macrophages. J Immunol. 2017;199(10):3614–22.

31. Pruim RJ, Welch RP, Sanna S, Teslovich TM, Chines PS, Gliedt TP, et al. LocusZoom: regional visualization of genome-wide association scan results. Bioinformatics. 2010;26(18):2336–7.

32. Aguet F, Brown AA, Castel SE, Davis JR, He Y, Jo B, et al. Genetic effects on gene expression across human tissues. Nature. 2017;550(7675):204–13.

33. Wang Y, Yang Q, Shen S, Zhang L, Xiang Y, Weng X. Mst1 promotes mitochondrial dysfunction and apoptosis in oxidative stress-induced rheumatoid arthritis synoviocytes. Aging (Albany NY). 2020;12(16):16211–23.

34. Zhang J, Sun L, Li W, Wang Y, Li X, Liu Y. Overexpression of macrophage stimulating 1 enhances the anti-tumor effects of IL-24 in esophageal cancer via inhibiting ERK-Mfn2 signaling-dependent mitophagy. Biomed Pharmacother. 2019;114:108844.

35. Freshour SL, Kiwala S, Cotto KC, Coffman AC, McMichael JF, Song JJ, et al. Integration of the Drug–Gene Interaction Database (DGIdb 4.0) with open crowdsource efforts. Nucleic Acids Research. 2020;49(D1):D1144–D51.

36. Chai YC, Hendrich S, Thomas JA. Protein S-thiolation in hepatocytes stimulated by t-butyl hydroperoxide, menadione, and neutrophils. Arch Biochem Biophys. 1994;310(1):264–72.

37. Mallis RJ, Hamann MJ, Zhao W, Zhang T, Hendrich S, Thomas JA. Irreversible thiol oxidation in carbonic anhydrase III: protection by S-glutathiolation and detection in aging rats. Biol Chem. 2002;383(3-4):649–62.

38. Li D, Burch P, Gonzalez O, Kashork CD, Shaffer LG, Bachinski LL, et al. Molecular cloning, expression analysis, and chromosome mapping of WDR6, a novel human WD-repeat gene. Biochem Biophys Res Commun. 2000;274(1):117–23.

39. Sivan G, Ormanoglu P, Buehler EC, Martin SE, Moss B. Identification of Restriction Factors by Human Genome-Wide RNA Interference Screening of Viral Host Range Mutants Exemplified by Discovery of SAMD9 and WDR6 as Inhibitors of the Vaccinia Virus K1L-C7L-Mutant. mBio. 2015;6(4):e01122.

40. Smith TF. Diversity of WD-Repeat proteins. In: Clemen CS, Eichinger L, Rybakin V, editors. The Coronin Family of Proteins: Subcellular Biochemistry. New York, NY: Springer New York; 2008. p. 20–30.

41. Chiba T, Inoue D, Mizuno A, Komatsu T, Fujita S, Kubota H, et al. Identification and characterization of an insulin receptor substrate 4-interacting protein in rat brain: implications for longevity. Neurobiol Aging. 2009;30(3):474–82.

42. McKnight NC, Jefferies HB, Alemu EA, Saunders RE, Howell M, Johansen T, et al. Genome-wide siRNA screen reveals amino acid starvation-induced autophagy requires SCOC and WAC. Embo j. 2012;31(8):1931–46.

43. Frendo-Cumbo S, Tokarz VL, Bilan PJ, Brumell JH, Klip A. Communication Between Autophagy and Insulin Action: At the Crux of Insulin Action-Insulin Resistance? Front Cell Dev Biol. 2021;9:708431.

44. Rautou PE, Cazals-Hatem D, Moreau R, Francoz C, Feldmann G, Lebrec D, et al. Acute liver cell damage in patients with anorexia nervosa: a possible role of starvation-induced hepatocyte autophagy. Gastroenterology. 2008;135(3):840–8, 8.e1-3.

45. van Niekerk G, Loos B, Nell T, Engelbrecht AM. Autophagy--A free meal in sickness-associated anorexia. Autophagy. 2016;12(4):727–34.

46. Adams DM, Reay WR, Geaghan MP, Cairns MJ. Investigation of glycaemic traits in psychiatric disorders using Mendelian randomisation revealed a causal relationship with anorexia nervosa. Neuropsychopharmacology. 2021;46(6):1093–102.

47. Reay WR, Kiltschewskij DJ, Geaghan MP, Atkins JR, Carr VJ, Green MJ, et al. Genetic estimates of correlation and causality between blood-based biomarkers and psychiatric disorders. Sci Adv. 2022;8(14):eabj8969.

48. Dalton B, Bartholdy S, Robinson L, Solmi M, Ibrahim MAA, Breen G, et al. A meta-analysis of cytokine concentrations in eating disorders. J Psychiatr Res. 2018;103:252–64.

49. Gibson D, Mehler PS. Anorexia Nervosa and the Immune System-A Narrative Review. J Clin Med. 2019;8(11).

50. Chanda D, Li J, Oligschlaeger Y, Jeurissen ML, Houben T, Walenbergh SM, et al. MSP is a negative regulator of inflammation and lipogenesis in ex vivo models of non-alcoholic steatohepatitis. Exp Mol Med. 2016;48(9):e258.

51. Lu K, Zhao J, Liu W. Macrophage stimulating 1-induced inflammation response promotes aortic aneurysm formation through triggering endothelial cells death and activating the NF-κB signaling pathway. J Recept Signal Transduct Res. 2020;40(4):374–82.

52. Kong D, Shen Y, Liu G, Zuo S, Ji Y, Lu A, et al. PKA regulatory IIα subunit is essential for PGD2-mediated resolution of inflammation. J Exp Med. 2016;213(10):2209–26.

53. Dalton B, Campbell IC, Chung R, Breen G, Schmidt U, Himmerich H. Inflammatory Markers in Anorexia Nervosa: An Exploratory Study. Nutrients. 2018;10(11).

54. Caso JR, Graell M, Navalón A, MacDowell KS, Gutiérrez S, Soto M, et al. Dysfunction of inflammatory pathways in adolescent female patients with anorexia nervosa. Progress in Neuro-Psychopharmacology and Biological Psychiatry. 2020;96:109727.

55. Del Giudice M, Gangestad SW. Rethinking IL-6 and CRP: Why they are more than inflammatory biomarkers, and why it matters. Brain Behav Immun. 2018;70:61–75.

